# Apathy in Mild Behavioural Impairment: Associations with Cortical Thickness and Grey Matter Volume

**DOI:** 10.64898/2026.02.25.26347107

**Authors:** Daniella Vellone, Dylan X. Guan, Zahra Goodarzi, Nils D. Forkert, Eric E. Smith, Zahinoor Ismail

**Affiliations:** Faculty of Graduate Studies, University of Calgary, Calgary, Alberta T2N 1N4, Canada; Hotchkiss Brain Institute, Cumming School of Medicine, University of Calgary, Calgary, Alberta T2N 4N1, Canada; Mathison Centre for Mental Health Research and Education, Cumming School of Medicine, University of Calgary, Calgary, Alberta T2N 4Z6, Canada; Department of Community Health Sciences, Cumming School of Medicine, University of Calgary, Calgary, Alberta T2N 4N1, Canada; Department of Medicine, Cumming School of Medicine, University of Calgary, Calgary, Alberta T2N 4N1, Canada; O’Brien Institute for Public Health, Cumming School of Medicine, University of Calgary, Calgary, Alberta T2N 4Z6, Canada; Department of Clinical Neurosciences, Cumming School of Medicine, University of Calgary, Calgary, Alberta T2N 4N1, Canada; Department of Radiology, Cumming School of Medicine, University of Calgary, Calgary, Alberta T2N 4N1, Canada; Alberta Children’s Hospital Research Institute, University of Calgary, Calgary, Alberta T3B 6A8, Canada; Department of Psychiatry, Cumming School of Medicine, University of Calgary, Calgary, Alberta T2N 4N1, Canada; Department of Pathology and Laboratory Medicine, University of Calgary, Calgary, Alberta T2N 4N1, Canada; Clinical and Biomedical Sciences, Faculty of Health and Life Sciences, University of Exeter, Exeter EX1 2HZ, United Kingdom

**Keywords:** Frontal, Motivation, Neuroimaging, Preclinical, Prodromal, Structural

## Abstract

Mild Behavioural Impairment (MBI) is defined by later-life onset of persistent behavioural changes and is recognized as a risk marker for cognitive decline and dementia. Apathy, a core MBI domain characterized by diminished interest, initiative, and emotional reactivity, can emerge before dementia and is hypothesized to be associated with structural brain changes.

While previous studies have explored Alzheimer disease (AD)-related neuroanatomical substrates of apathy in the dementia clinical stage, few have investigated these associations in cognitively normal (CN) or mild cognitive impairment (MCI) individuals with persistent apathy consistent with MBI. Thus, this study explores structural brain differences between individuals with MBI-apathy and those without neuropsychiatric symptoms (no-NPS).

Participants (n = 446; mean age = 69.6 years; 79.8% CN; 62.8% female) were drawn from the National Alzheimer’s Coordinating Center and categorized into MBI-apathy (n = 59) and no-NPS (n = 387) groups. Linear regressions were used to model associations between NPS group and regional brain measures, with adjustments for age, sex, years of education, apolipoprotein E4 carrier status, intracranial volume, and Mini-Mental State Examination score, with false discovery rate (FDR) correction for multiple comparisons. Primary outcomes included two predefined AD meta-regions-of-interest (ROIs): 1) thickness: a composite measure of mean cortical thickness across the entorhinal cortex, inferior temporal gyrus, middle temporal gyrus, inferior parietal lobule, fusiform gyrus, and precuneus; and 2) volume: a composite measure of mean cortical and subcortical grey matter volume across the hippocampus, entorhinal cortex, amygdala, middle temporal gyrus, inferior parietal lobule, and precuneus. Primary outcomes also included cortical thickness and grey matter volume among individual ROIs including the ventral striatum (VS), anterior cingulate cortex (ACC), orbitofrontal cortex (OFC), ventrolateral prefrontal cortex (vlPFC), and dorsolateral prefrontal cortex (dlPFC).

MBI-apathy status was associated with significantly lower AD-meta-ROI cortical thickness (*Z*-score difference [95% CI]; FDR-corrected *p*-value, -0.43 [-0.73 – [-0.12]]; 0.025) and lower AD meta-ROI grey matter volume (-0.50 [-0.71 – [-0.30]]; <0.001). MBI-apathy was also associated with significantly lower dlPFC thickness (-0.40, [-0.70 – [-0.09]]; 0.02) and volume (-0.28 [-0.50– [-0.06]]; 0.026) and lower OFC volume (-0.32, [-0.57 – [-0.07]]; 0.026) compared to the no-NPS group.

Within a non-dementia sample, MBI-apathy was more strongly associated with established AD-vulnerable regions than with regions that have been traditionally implicated in apathy in dementia. Results suggests that during CN and MCI stages, MBI-apathy may reflect early AD-related neurodegeneration, with conventional apathy-related structural changes becoming more prominent as disease progresses.

## Introduction

Apathy is among the most common neuropsychiatric symptoms (NPS) in dementia, affecting over half of affected individuals.^1^ However, its clinical significance is not limited to dementia. Apathy can emerge and persist at earlier disease stages along the clinical continuum, with prevalence estimates of 14% in cognitively normal (CN) individuals and 29% in mild cognitive impairment (MCI).^2^ Characterized by a marked reduction in interest, goal-directed behaviour, and emotional responsiveness, apathy occurs independently of motor or sensory impairments and can significantly hinder an individual’s ability to engage in daily activities and maintain meaningful social relationships.^3–6^ While apathy is well documented in Alzheimer disease (AD) dementia, particularly in the context of behavioural and psychological symptoms of dementia (BPSD), there is less research on apathy in CN individuals and those with MCI.

The emergence and persistence of apathy in non-dementia stages is captured by the mild behavioural impairment (MBI) framework, a validated neurobehavioural syndrome that describes the later-life onset of persistent behaviour or personality changes as an at-risk state for cognitive decline and incident dementia.^7^ MBI and BPSD lie on a continuum, with MBI serving as a critical nosological bridge between NPS and neurodegeneration. Conceptually, this framework offers a clinically useful lens that enables the investigation of early changes in brain-behaviour relationships in a consistent, specific, and validated manner.^7–12^ Our previous research demonstrated that dementia-free individuals who exhibit emergent and persistent apathy meeting MBI criteria (MBI-apathy) are significantly more likely to progress to dementia, with approximately 81% of progressors eventually developing AD dementia.^13^ Additionally, MBI-apathy has been associated with core AD biomarkers, including cerebrospinal fluid (CSF) p-tau_181_, p-tau_181_/Aβ_42_, t-tau, and t-tau/Aβ_42_, both cross-sectionally and over two years.^14^ In contrast, individuals with NPS not meeting MBI criteria (non-MBI NPS) did not show significant associations compared to those without NPS. MBI-apathy was also significantly associated with plasma p-tau_181_ at baseline, and over two- and three-year time points, whereas those with non-MBI NPS did not exhibit these associations.^15^

Despite these findings, the link between MBI-apathy and structural brain changes remains largely unexplored, particularly in the context of early-stage neurodegeneration. In AD dementia, apathy has been associated with structural brain changes in regions traditionally linked to cognition, including those in the temporal lobe.^16,17^ Beyond these AD-related brain regions, neuroimaging studies have demonstrated that apathy is associated with dysfunction in brain regions involved in emotional regulation and goal-directed behaviour, particularly the ventral striatum (VS), anterior cingulate cortex (ACC), orbitofrontal cortex (OFC), ventrolateral prefrontal cortex (vlPFC), and dorsolateral prefrontal cortex (dlPFC).^18–20^ These brain regions are central to the frontal-subcortical circuits that mediate motivation and executive function, with atrophy in these areas in AD dementia patients consistently linked to apathy.^21^ Recent studies also suggest that amyloid and tau deposition in these regions may occur independent of cognitive decline and contribute to the onset of apathy in preclinical and prodromal stages.^18,19^ Together, these findings suggest that apathy in AD-dementia results from widespread neurodegenerative changes in both temporal and frontal-subcortical circuits, consistent with fMRI findings in non-dementia samples.^22^

Despite these insights, studies exploring apathy-related brain changes in the early stages of disease are limited. Much of the existing neuroimaging literature has focused on individuals with AD dementia, implicitly positioning the regions most affected in dementia as the primary neural substrates of apathy. However, apathy frequently emerges and persists during preclinical and prodromal stages, when many of these frontal and temporal regions may not yet exhibit marked structural change. While the presence of apathy in these early disease stages has been recognized as a potential marker of future dementia,^13,23^ the specific brain regions associated with apathy prior to dementia onset, therefore, remain less clear.

In this study, we investigate the structural brain changes associated with MBI-apathy in CN and MCI individuals. We hypothesized that MBI-apathy would be associated with lower cortical thickness and lower cortical and subcortical grey matter volume in regions typically implicated in AD and motivation, compared to individuals without NPS (no-NPS). Specifically, we expected associations with composite AD meta-regions of interest (meta-ROIs), which summarize structural differences across multiple AD-vulnerable regions, as well as with apathy-related regions including the VS, ACC, OFC, vlPFC, and dlPFC. By investigating structural brain differences between individuals with MBI-apathy and those without NPS, this study aims to provide a better understanding of the neurobiological substrates underlying apathy during preclinical and prodromal stages of AD.

## Materials and Methods

### Study Population: National Alzheimer’s Coordinating Centre (NACC)

Participant data for this cross-sectional observational study were drawn from the National Alzheimer’s Coordinating Center (NACC; https://naccdata.org/), a large multi-center dataset that aggregates longitudinal evaluations from National Institute on Aging (NIA)-funded Alzheimer’s Disease Research Centers (ADRCs) across the United States.^24,25^ Visits were conducted approximately annually, during which extensive data were collected, including detailed clinical assessments (*e.g.,* demographics, family and medical history, physical and neurological exams, and diagnostic evaluations), cognitive testing, neuropsychiatric evaluations, and measures of functional status (*e.g.,* activities of daily living). In addition, many participants contributed neuroimaging data (*e.g.,* MRI scans). The current study utilized data from 45 ADRCs, encompassing participant visits conducted between June 2005 and February 2022. Although the NACC cohort spans the entire cognitive spectrum,^26^ from CN individuals to those diagnosed with dementia, only participants without dementia, at CN and MCI stages, were included in this analysis. All ADRCs obtained ethics approval from their respective institutions prior to submitting data to NACC, and all procedures at participating ADRCs were conducted in accordance with the principles of the Declaration of Helsinki. Detailed descriptions of NACC recruitment and data collection procedures can be found elsewhere.^24,25,27^

### Participant Selection

A study participant selection flow diagram for data analysis is shown in Fig. 1. Participants were included in this study if they had complete Neuropsychiatric Inventory Questionnaire (NPI-Q)^28^ domain scores necessary for determining NPS status, and at least two study visits within the first two years to determine symptom persistence.

**Figure 1.**
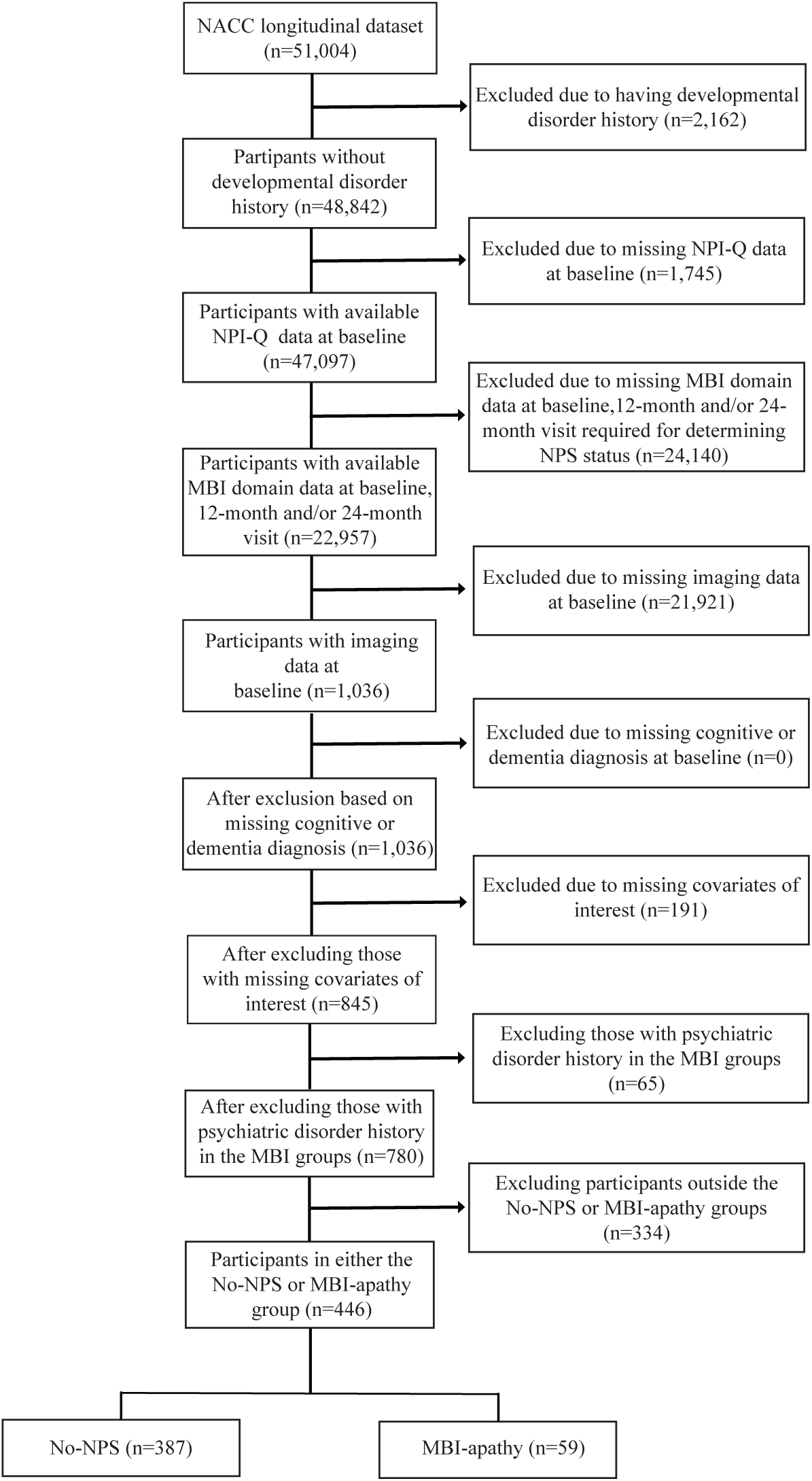
Flowchart of participants from NACC included for analysis. CN: Cognitively Normal, MCI: Mild Cognitive Impairment, MBI: Mild Behavioural Impairment, NPI-Q: Neuropsychiatric Inventory Questionnaire, NPS: Neuropsychiatric Symptoms.

### MBI Operationalization

To determine MBI domain scores, NPI-Q scores were transformed into MBI domain scores using a published and validated algorithm.^29^ In this study, we operationalized MBI-apathy using the NPI-Q apathy domain scores.^30^ The NPI-Q assesses apathy using a single question that distills key aspects of the NPI apathy screening item, focusing on diminished interest in both personal activities and social engagement.^28^ The presence of apathy was determined by an apathy domain score > 0. To meet the MBI-apathy symptom persistence criterion,^29^ apathy needed to be identified in at least two of three visits within the first two years, either at baseline and 12 months, baseline and 24 months, 12 and 24 months, or 0, 12, and 24 months. Individuals were included in the MBI-apathy group irrespective of whether they had concurrent NPS in other MBI domains. The no-NPS group included participants without any NPS (MBI total score = 0) at all visits up to two years.

### MRI Data Acquisition and Processing

All T1-weighted structural MRI brain scans were processed using FreeSurfer version 6.0 image analysis suite,^31,32^ executed on the Canadian Brain Imaging Research Platform (CBRAIN),^33^ a web-based high-performance computing environment for neuroimaging analyses. The standard FreeSurfer *recon-all* pipeline was applied to each scan, involving multiple automated steps, including motion correction, spatial normalization (*i.e.,* Talairach registration), intensity normalization, skull stripping, and tissue segmentation with anatomical labeling of cortical and subcortical brain structures.^31^ This pipeline outputs regional cortical thickness and cortical grey matter volume across 34 bilateral cortical regions defined by the Desikan-Killiany atlas,^34^ as well as grey matter volumes for 18 bilateral subcortical structures.^32^

After processing, all images were visually inspected for structural abnormalities, imaging artifacts, and segmentation errors. Scans with identifiable repairable issues were manually edited and reprocessed, while those with irreparable errors were excluded from the analysis.

To account for variability in imaging protocols across participating sites and differences in scanner characteristics, we applied the Normative Morphometry Image Statistics (NOMIS) tool (https://github.com/medicslab/NOMIS).^12^ NOMIS provides a normative reference model derived from about 7000 cognitively healthy adults (aged 18-100 years) and includes over 1000 brain morphometric measures computed by FreeSurfer. For each cortical thickness and grey matter volume measure, NOMIS generates a standardized *Z*-score indicating the extent to which that value deviates from the expected norm (where the population mean is 0 and the standard deviation is 1). NOMIS inherently adjusts for image quality factors (*e.g.,* voxel size, contrast-to-noise ratio, surface reconstruction holes) and participant characteristics (*e.g.,* age, sex, intracranial volume).

Analyses were restricted to *a priori* selected cortical and subcortical regions relevant to AD and apathy, rather than the full set of FreeSurfer-derived regions. To capture AD-related neurodegeneration, we employed previously defined AD “signature” meta-ROIs, which summarize structural change across several brain regions known to be affected early in the AD course.^35^ Consistent with prior work,^35^ the AD cortical thickness meta-ROI was calculated as the mean of regional cortical thickness *Z*-scores across the entorhinal cortex, inferior temporal lobe, middle temporal lobe, inferior parietal lobe, fusiform gyrus, and precuneus. For each region, left and right hemisphere values first were averaged to derive bilateral measures before computing regional *Z*-scores, which were then averaged to generate a single composite cortical thickness *Z*-score. Although prior work summed regional volumes,^35^ we used an averaged *Z*-score composite to maintain consistency with the cortical thickness meta-ROI and to facilitate interpretability of standardized effect sizes. Accordingly, the AD volume meta-ROI was calculated as the mean of regional grey matter volume *Z*-scores across the hippocampus, entorhinal cortex, amygdala, middle temporal lobe, inferior parietal lobe, and precuneus, using the same bilateral averaging approach.^35^ In parallel, apathy-related ROIs were selected *a priori* to capture frontal-subcortical circuits most consistently implicated in motivational and goal-directed behaviour in the neuroimaging literature.^18–20^ The VS (volume only), ACC, and OFC were derived directly from Desikan-Killiany parcels. The dlPFC and vlPFC were constructed as composite regions based on anatomically relevant Desikan–Killiany parcels. Specifically, the dlPFC was calculated as the mean of bilateral caudal middle frontal, rostral middle frontal, and superior frontal *Z*-scores. The vlPFC was calculated as the mean of bilateral pars opercularis, pars orbitalis, and pars triangularis *Z*-scores. For each composite, left and right hemisphere values were first averaged to derive bilateral measures prior to computing regional *Z*-scores.^36^

### Statistical Analysis

We examined baseline demographic characteristics, including age, sex, and years of education. Clinical variables included Mini-Mental State Examination (MMSE) score and NPI-Q. Biomarker data included apolipoprotein E4 (*APOE4*) carrier status, as well as *Z*-scores for cortical thickness and cortical and subcortical grey matter volume in the VS, ACC, OFC, vlPFC, dlPFC, and the AD cortical thickness and AD grey matter volume meta-ROIs. The MBI-apathy group was compared to the no-NPS group using χ^2^ tests for categorical variables and independent samples *t*-tests for continuous variables.

Linear regression models controlled for years of education, *APOE4* carrier status, and MMSE score (with the NOMIS tool adjusting for other variables). NPS status was modeled as a categorical predictor with two levels, including MBI-apathy and no-NPS, with the no-NPS group serving as the reference. *APOE4* carrier status was similarly modeled as a binary variable, where individuals with one or more *APOE4* alleles were classified as *APOE4* carriers, and those with no *APOE4* alleles as non-*APOE4* carriers.

Statistical analyses were conducted in R v4.4.0, with the *stats* package for linear models. To account for multiple testing and control the false discovery rate (FDR), we applied the Benjamini-Hochberg correction procedure to the *p*-values obtained from our analyses, adjusting separately for five cortical thickness measures and six cortical and subcortical grey matter volume measures. Statistical significance was defined as an adjusted *p*-value < 0.05.

Assumptions for linear regression modeling were assessed and confirmed using diagnostic plots, which included Residuals versus Fitted, Normal Q-Q, Scale-Location, and Residuals versus Leverage to evaluate linearity, normality of errors, equal variance of errors, and the presence of influential data points.

### Data availability

The data that support the findings of this study are available on request from NACC (https://naccdata.org/requesting-data/data-request-process). Data cleaning and analysis scripts used in the present study are available from the authors upon reasonable request.

## Results

### Sample Composition and Demographic Profile

The study sample included in this work comprised 387 participants with no NPS and 59 participants with MBI-apathy. Average age was 69.6 years old, with 62.8% being female. Of the participants, 79.8% were CN and 20.2% had MCI. Compared to the no-NPS group, those with MBI-apathy were older, more likely to be male, and had a higher likelihood of being *APOE4* carriers. Those with MBI-apathy also exhibited lower MMSE scores and a higher prevalence of MCI (Table 1).

**Table 1.** Demographic and clinical characteristics by NPS group.

|  | No-NPS<br>(n=387) | MBI-apathy<br>(n=59) | *p*-value |
| --- | --- | --- | --- |
| **Age** |  |  |  |
| Mean (SD) | 68.9 (10.4) | 74.1 (7.62) | **<0.001<sup>a</sup>** |
| Median [Min, Max] | 68.0 [50.0, 92.0] | 75.0 [55.0, 89.0] |  |
| **Sex** |  |  |  |
| Male | 123 (31.8%) | 43 (72.9%) | **<0.001<sup>b</sup>** |
| Female | 264 (69.2%) | 16 (27.1%) |  |
| **Years of education** |  |  |  |
| Mean (SD) | 16.0 (2.89) | 15.1 (4.05) | 0.057a |
| Median [Min, Max] | 16.0 [5.00, 25.0] | 16.0 [2.00, 22.0] |  |
| **APOE4 Carrier Status** |  |  |  |
| Non-APOE4 Carrier | 240 (62.0%) | 23 (39.0%) | **0.001<sup>b</sup>** |
| APOE4 Carrier | 147 (38.0%) | 36 (61.0%) |  |
| **MMSE Score** |  |  |  |
| Mean (SD) | 28.9 (1.36) | 26.5 (2.58) | **<0.001<sup>a</sup>** |
| Median [Min, Max] | 29.0 [23.0, 30.0] | 27.0 [19.0, 30.0] |  |
| **Cognitive Status** |  |  |  |
| CN | 345 (89.1%) | 11 (18.6%) | **<0.001<sup>b</sup>** |
| MCI | 42 (10.9%) | 48 (81.4%) |  |

NPS = Neuropsychiatric Symptoms; MBI = Mild Behavioural Impairment; APOE = Apolipoprotein E; MMSE = Mini-Mental State Examination; CN = Cognitively Normal; MCI = Mild Cognitive Impairment; SD = Standard deviation.
Significant *p*-values are shown in bold.
<sup>a</sup>Independent samples *t*-test.
<sup>b</sup> $\chi^2$ tests.

### Group Differences and Cross-Sectional Associations Between MBI-Apathy and Brain Structure

Unadjusted descriptive group comparisons of regional brain measures, reported as *Z*-scores, are provided in Supplementary Table 1, with corresponding distributions shown in Fig. 2.

**Figure 2.**
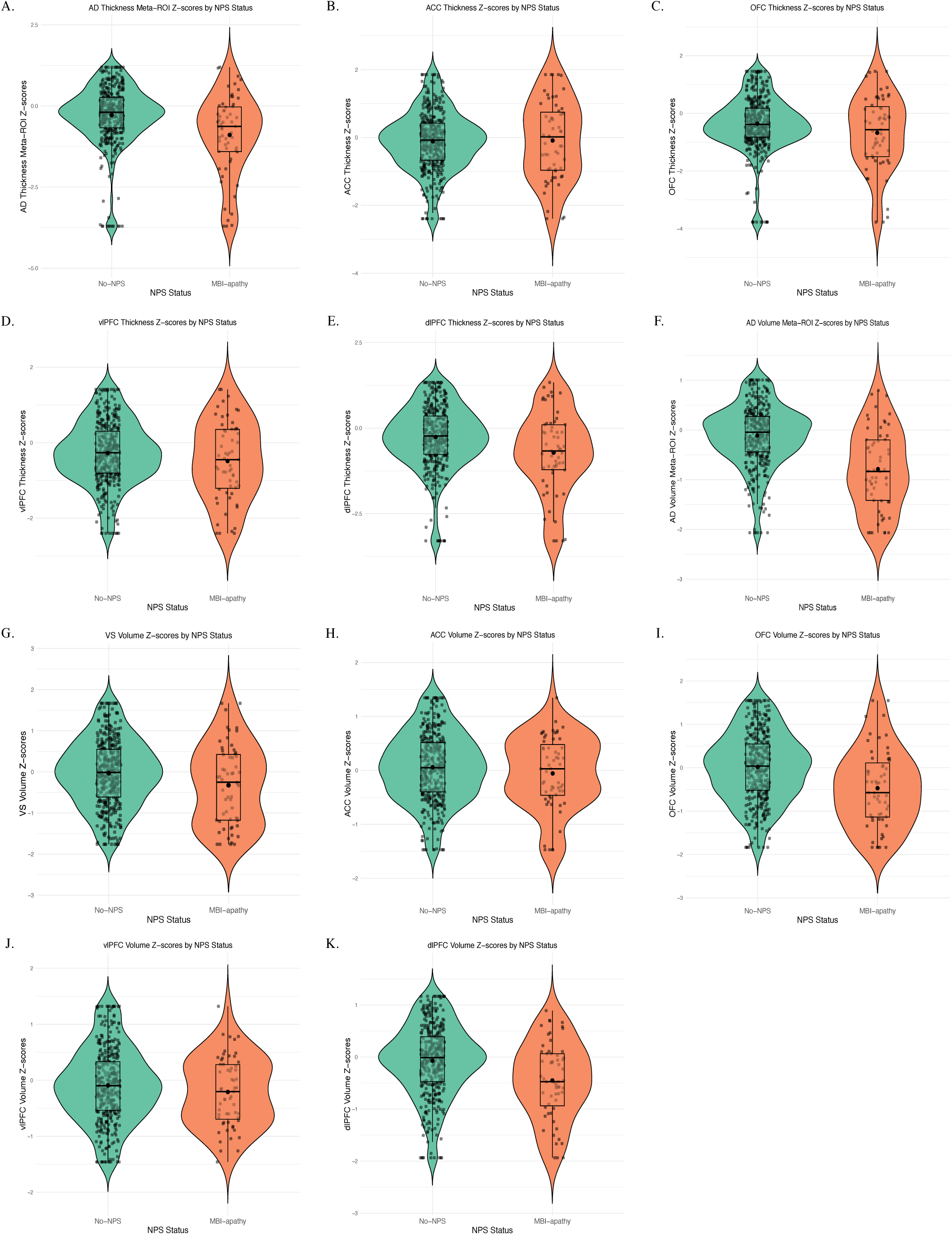
Violin plots showing the distribution of brain region differences by NPS group. Plots include jittered data points, boxplots representing the interquartile range and median, and mean values. NPS: Neuropsychiatric Symptoms; MBI: Mild Behavioural Impairment; Ventral striatum (VS); Anterior cingulate cortex (ACC); Orbitofrontal cortex (OFC); Ventrolateral prefrontal cortex (vlPFC); Dorsolateral prefrontal cortex (dlPFC); AD: Alzheimer’s Disease; ROI: Region of interest.

In adjusted linear regression models, compared to the no-NPS group, MBI-apathy was associated with significantly lower AD-signature composite *Z*-scores for both cortical thickness and cortical and subcortical grey matter volume (Table 2). Specifically, individuals with MBI-apathy exhibited 0.43 standard deviations lower AD thickness meta-ROI *Z*-scores (*Z*-score difference [95% CI]; FDR-corrected *p*-value, -0.43 [-0.73 – [-0.12]]; 0.025) and 0.50 standard deviations lower AD volume meta-ROI *Z*-scores (-0.50 [-0.71 – [-0.30]]; <0.001) compared to individuals without NPS.

**Table 2.** Adjusted associations of MBI-apathy with regional cortical thicknesses and grey matter volumes.

| Outcome | Z-score Difference | 95% CI | *p*-value | FDR-adjusted *p*-value |
| --- | --- | --- | --- | --- |
| AD thickness meta-ROI | -0.43 | -0.73 – [-0.12] | **0.006** | **0.025** |
| ACC thickness | 0.02 | -0.27 – 0.31 | 0.917 | 0.917 |
| OFC thickness | -0.28 | -0.60 – 0.03 | 0.079 | 0.132 |
| vIPFC thickness | -0.22 | -0.50 – 0.05 | 0.109 | 0.136 |
| dlPFC thickness | -0.40 | -0.70 – [-0.09] | **0.010** | **0.025** |
| AD volume meta-ROI | -0.50 | -0.71 – [-0.30] | **<0.001** | **<0.001** |
| VS volume | -0.26 | -0.53 – 0.00 | **0.050** | 0.075 |
| ACC volume | -0.07 | -0.27 – 0.14 | 0.529 | 0.529 |
| OFC volume | -0.32 | -0.57 – [-0.07] | **0.013** | **0.026** |
| vIPFC volume | -1.24 | -0.33 – 0.08 | 0.239 | 0.287 |
| dlPFC volume | -0.28 | -0.50 – [-0.06] | **0.012** | **0.026** |

Z-score differences represent the estimated differences in regional cortical thickness and grey matter volume Z-scores between the MBI-apathy and no-NPS groups, derived from linear regression models adjusted for years of education, APOE4 carrier status, and Mini-Mental State Examination score. Age, sex, and intracranial volume were accounted for during image preprocessing. NPS = Neuropsychiatric Symptoms; MBI = Mild Behavioural Impairment; VS = Ventral striatum; ACC = Anterior cingulate cortex; OFC = Orbitofrontal cortex; vIPFC = Ventrolateral prefrontal cortex; dlPFC = Dorsolateral prefrontal cortex; AD = Alzheimer's Disease; ROI = Region of interest; CI = Confidence interval; FDR = False Discovery Rate. Significant *p*-values are shown in bold.

Additionally, MBI-apathy status was associated with significantly lower cortical thickness and volume of the dlPFC and significantly lower volume of the OFC (Table 2). Specifically, individuals with MBI-apathy exhibited 0.40 standard deviations lower dlPFC thickness (-0.40, [-0.70 – [-0.09]]; 0.025), 0.28 standard deviations lower dlPFC volume (-0.28 [-0.50 – [-0.06]]; 0.026), and 0.32 standard deviations lower OFC volume (-0.32, [-0.57 – [-0.07]]; 0.026) compared to the no-NPS group. No significant associations were observed for VS volume (-0.26 [-0.53 – 0.00]; 0.075), ACC thickness (0.02 [-0.27 – 0.31]; 0.917) or volume (-0.07 [-0.27 – 0.14]; 0.529), OFC thickness (-0.28, [-0.60 – 0.03]; 0.132), and vlPFC thickness (-0.22 [-0.50 – 0.05]; 0.136) or volume (-1.24 [-0.33 – 0.08]; 0.287).

## Discussion

In this study of 446 dementia-free participants, we investigated cross-sectional associations between MBI-apathy and regional cortical thickness and grey matter volume in brain regions implicated in AD and motivational processes. Compared to individuals without NPS, MBI-apathy status was associated with significantly lower thickness and volume in AD-specific meta-ROIs, and significantly lower thickness and volume in the dlPFC and lower volume in the OFC. No significant differences were observed in the VS, ACC, or vlPFC. Overall, our findings suggest that MBI-apathy is associated with selective structural changes in AD-vulnerable and prefrontal regions prior to dementia onset.

Apathy has been associated with neurodegenerative changes in regions often affected in AD, with associations initially reported in dementia and more recently in CN and MCI individuals.^37,38^ Structural MRI studies of patients with AD dementia have reported reduced cortical thickness in temporal regions, including the temporal cortex and temporal pole.^17,37^ Functional MRI findings have similarly implicated medial temporal-limbic structures, with altered activity in the amygdala among individuals with apathy.^39^ Single-photon emission computed tomography (SPECT) studies demonstrated reduced perfusion across temporal cortical regions implicated in AD, including the amygdala and areas of the superior and inferior temporal cortex.^40,41^ Positron emission tomography (PET) studies further showed that higher baseline entorhinal tau burden is associated with greater apathy severity over time,^42^ linking apathy to AD-related pathological processes in medial temporal networks. In cognitively unimpaired and MCI samples, lower inferior temporal cortical thickness and elevated amyloid-β and entorhinal tau burden have been associated with greater apathy.^43,44^ Further, neuroimaging studies incorporating the MBI construct reported associations between global MBI and AD-related regions, including reduced hippocampal and entorhinal volumes and lower AD meta-ROI cortical thickness,^12,36^ alongside greater medial temporal tau signal and temporal hypoperfusion.^45,46^ To our knowledge, only two MBI studies have examined AD-related grey matter brain structural correlates by behavioural domain, one reporting an association between hippocampal volume and apathy and the other reporting associations between the posterior cingulate and precuneus with apathy.^36,47^ However, no prior work has specifically examined cortical thickness and cortical and subcortical grey matter volume correlates of MBI-apathy in individuals without dementia.

In the present study, MBI-apathy was strongly associated with lower cortical thickness and lower cortical and subcortical volume in AD-related regions. Associations were particularly pronounced in AD meta-ROIs encompassing the hippocampus, entorhinal cortex, amygdala, inferior and middle temporal lobe, inferior parietal lobe, fusiform gyrus, and precuneus; brain areas that are known to be among the earliest affected by AD pathology.^35^ The 0.50-SD difference in AD volume meta-ROI *Z*-scores between MBI-apathy and no-NPS groups highlights the degree to which apathy may signal emerging AD-related neurodegeneration. Alignment with prior work further strengthens this interpretation, particularly studies showing that global MBI scores are associated with atrophy in medial temporal regions even among CN older adults.^12,36^ The association of MBI-apathy with lower cortical thickness and volume in AD meta-ROIs is also consistent with Braak staging of tau pathology. Early stages (Braak I-II) show changes centred in the transentorhinal/entorhinal cortex, intermediate stages (Braak III-IV) extend into the hippocampus and adjacent medial temporal association cortex (*e.g.,* fusiform, medial temporal gyrus), and late stages (Braak V-VI) reflect diffuse neocortical involvement.^48,49^ Therefore, the atrophy of entorhinal, medial temporal, and temporoparietal regions in persons with apathy in our sample of CN/MCI likely reflects early-to-intermediate stage AD (Braak stages I-IV) rather than late-stage widespread neocortical involvement. This staging-concordant topography strengthens the interpretation that MBI-apathy reflects early AD-related network vulnerability, emerging along with the canonical trajectory of tau spread before widespread cortical degeneration and clinical dementia. In this context, MBI-apathy may be related to a disruption of the medial temporal and temporoparietal circuits at a stage when cognitive impairment is not yet clinically apparent, aligning with models that position NPS as an early behavioural manifestation of AD pathophysiology.

Beyond AD-related regions, structural correlates of apathy in AD dementia have been documented, where widespread atrophy in frontolimbic and frontal-subcortical circuits contribute to motivational impairment.^18,20^ Previous structural MRI studies have consistently identified associations between apathy and lower volume and thickness in the ACC, posterior cingulate cortex, OFC, dlPFC, parts of the basal ganglia, and insula,^50–57^ suggesting dysfunction in behavioural initiation, executive control, and reward valuation.^58^ Grey matter structural MRI findings are complimented by converging evidence from other imaging modalities, including white matter structural MRI, ^43,59–63^ functional MRI,^39,64–66^ SPECT,^40,41,67–73^ and PET.^42,74–81^ However, most previous research has focused on individuals with dementia, where neuropathological burden is already significant, and studies examining apathy in non-dementia samples frequently combine MCI with dementia in the analyses.^16,42,60^ Since dementia cases typically exhibit pronounced neurodegeneration, effect estimates in such mixed samples are often driven by later-stage changes. This can mask subtler structural alterations that emerge during prodromal stages and limit insight into early disease mechanisms. Among studies that specifically examined non-dementia samples, apathy in some studies has been associated with abnormalities in the ACC, OFC, dlPFC, and areas of the basal ganglia and limbic system.^16,42,82–85^ However, other investigations have reported no significant neuroanatomical or metabolic correlates of apathy during these earlier stages,^38,86–88^ potentially reflecting variability in how apathy has been operationalized across studies, highlighting the need for further research specifically focussed in non-dementia cohorts. Taken together, the findings suggest that apathy emerging before dementia may signal early neurobiological vulnerability. Within this context, the MBI framework characterizes persistent later-life behavioural change, offering a structured means to interrogate early neural alterations associated with apathy. Relatedly, although neuroimaging studies of MBI remain limited, available evidence also supports involvement of frontal and paralimbic circuits with later-life emergent behavioural change. Previous structural MRI analyses have shown that higher MBI severity is associated with lower grey matter in frontal regions,^89,90^ including the ACC, OFC, middle and superior frontal gyri, as well as thalamic and insular regions,^90–92^ with additional reports of altered frontal-parietal connectivity and reduced frontal perfusion.^46,93^ However, these studies largely examined total MBI severity, which aggregates heterogeneous behavioural domains and therefore limits inferences about apathy-specific neuroanatomical substrates within MBI. The present study directly addresses this gap by focusing on individuals with apathy meeting MBI criteria to identify structural correlates of motivational symptoms prior to dementia.

The results of the present study align with prior work implicating the dlPFC and OFC in motivational processes. More precisely, MBI-apathy was associated with significantly lower dlPFC thickness and volume and OFC volume, suggesting that structural disruptions in lateral prefrontal circuits may underlie emergent and persistent apathy in older adults without dementia. The dlPFC is involved in executive functioning, planning, and goal maintenance, while the OFC plays a critical role in affective evaluation and reward-based decision-making.^94^ Dysfunction in these brain regions has been proposed to interfere with the formation and execution of goal-directed actions and with assigning motivational value to behaviour, consistent with diminished initiative and blunted emotional engagement observed in apathy.^94^ Our findings also support this mechanistic model in a non-dementia context, suggesting that prefrontal structural changes may be detectable prior to significant cognitive impairment. While prefrontal effects were robust, associations with brain regions traditionally implicated in apathy, including the VS and ACC, did not reach statistical significance, although directions of effect were consistent with prior literature.^16^ One potential explanation for this finding is that medial prefrontal and subcortical involvement becomes more prominent with advancing neurodegeneration, whereas lateral prefrontal changes may emerge relatively earlier. This interpretation aligns with PET findings, which show that apathy in preclinical and prodromal stages correlates more strongly with hypometabolism and tau burden in lateral prefrontal and temporoparietal regions, while medial prefrontal and subcortical changes become more pronounced in the dementia stage.^42,95^

### Limitations

Several limitations of this study should be acknowledged. First, apathy was measured using the NPI-Q, from which MBI-apathy was operationalized. Although the NPI-Q to MBI algorithm is validated and enables the application of MBI criteria in large observational cohorts, the NPI-Q includes a single apathy screening item and does not capture the full multidimensionality of apathy, including initiative, interest, and emotional reactivity subdomains.^5,30^ The MBI-Checklist provides a more comprehensive assessment of later-life emergent behavioural change, from which all three domains of apathy can be identified, the use of which would strengthen future studies.^8^ Second, although efforts were made to control for demographic and cognitive variables, residual confounding remains possible. Apathy frequently co-occurs with other NPS, such as agitation,^96,97^ which share overlapping mechanisms and may influence both behaviour and brain structure.^18^ Additionally, unmeasured psychosocial and environmental factors, such as social engagement, physical activity, or stress, may contribute to behavioural expression and neural vulnerability.^98–101^ Third, the cross-sectional design precludes inferences regarding directionality. Whether apathy reflects early neurodegenerative changes or contributes to accelerated decline cannot be determined; longitudinal neuroimaging analyses will be necessary to clarify temporal dynamics and disease progression. Fourth, participants were recruited through ADRCs and are generally highly educated, research-engaged, and less racially and socioeconomically diverse than the broader population. As such, generalizability to community-dwelling older adults may be limited, underscoring the need to examine MBI-apathy in more diverse and representative cohorts. Fifth, MRI data were collected across multiple sites using heterogeneous scanner types and acquisition parameters. While the use of the NOMIS normative harmonization framework mitigates scanner- and protocol-related variance to some extent, multi-site variability cannot be fully eliminated. Additionally, not all NACC participants contribute imaging data, but those who do may represent a healthier and more motivated subset, introducing potential selection bias.

Finally, our analyses focused on *a priori* ROIs informed by existing AD and apathy literature. Although theoretically grounded, this targeted approach may overlook structural differences in other cortical or subcortical regions or network-level alterations detectable through whole-brain or connectome-based analyses.

Future studies should build on this work by examining longitudinal changes in MBI-apathy and brain structure, incorporate biomarker-confirmed AD pathology, and expand to larger and more diverse samples. Additionally, multimodal imaging studies incorporating functional and structural connectivity measures may offer further insight into network-level disruptions underlying apathy in preclinical and prodromal stages.

## Conclusion

In this cohort of dementia-free older adults, MBI-apathy was associated with lower cortical thickness and lower cortical and subcortical volume in both AD-vulnerable and lateral prefrontal brain regions compared to those without NPS. These findings demonstrate that apathy, when defined within the MBI framework, maps onto neuroanatomical changes consistent with early AD neurodegeneration, even prior to clinically evident dementia. This pattern indicates that at the CN and MCI stages, MBI-apathy may reflect emerging associations with brain changes characteristic of early AD. Apathy-related regions traditionally implicated in dementia, such as medial frontal and subcortical circuits, may become more prominently affected later in the disease course, with stronger associations likely to emerge as individuals progress to dementia, supporting the conceptual bridge between MBI and BPSD.

Taken together, these findings highlight MBI-apathy as a clinically observable behavioural marker of early disease vulnerability and underscore the importance of incorporating neuropsychiatric changes into models of preclinical and prodromal AD. Therefore, identification of MBI-apathy may improve early risk detection and inform early intervention, enabling clinicians to recognize individuals at heightened vulnerability and consider increased monitoring, lifestyle and behavioural supports, or referral for biomarker assessment. Such proactive identification may ultimately enhance opportunities for person-centred planning, early therapeutic engagement, and future disease-modifying strategies as they become available.

## Data Availability

The redistribution of data obtained from NACC is restricted under the NACC Data Use Agreement (https://files.alz.washington.edu/documentation/nacc_data_use_agreement.pdf). Researchers interested in accessing NACC data can submit a request using the following link: https://naccdata.org/requesting-data/data-request-process. Data cleaning and analysis scripts used in the present study are available from the authors upon reasonable request.

## Acknowledgements

The NACC database is funded by NIA/NIH Grant U24 AG072122. NACC data are contributed by the NIA-funded ADRCs: P30 AG062429 (PI James Brewer, MD, PhD), P30 AG066468 (PI Oscar Lopez, MD), P30 AG062421 (PI Bradley Hyman, MD, PhD), P30 AG066509 (PI Thomas Grabowski, MD), P30 AG066514 (PI Mary Sano, PhD), P30 AG066530 (PI Helena Chui, MD), P30 AG066507 (PI Marilyn Albert, PhD), P30 AG066444 (PI David Holtzman, MD), P30 AG066518 (PI Lisa Silbert, MD, MCR), P30 AG066512 (PI Thomas Wisniewski, MD), P30 AG066462 (PI Scott Small, MD), P30 AG072979 (PI David Wolk, MD), P30 AG072972 (PI Charles DeCarli, MD), P30 AG072976 (PI Andrew Saykin, PsyD), P30 AG072975 (PI Julie A. Schneider, MD, MS), P30 AG072978 (PI Ann McKee, MD), P30 AG072977 (PI Robert Vassar, PhD), P30 AG066519 (PI Frank LaFerla, PhD), P30 AG062677 (PI Ronald Petersen, MD, PhD), P30 AG079280 (PI Jessica Langbaum, PhD), P30 AG062422 (PI Gil Rabinovici, MD), P30 AG066511 (PI Allan Levey, MD, PhD), P30 AG072946 (PI Linda Van Eldik, PhD), P30 AG062715 (PI Sanjay Asthana, MD, FRCP), P30 AG072973 (PI Russell Swerdlow, MD), P30 AG066506 (PI Glenn Smith, PhD, ABPP), P30 AG066508 (PI Stephen Strittmatter, MD, PhD), P30 AG066515 (PI Victor Henderson, MD, MS), P30 AG072947 (PI Suzanne Craft, PhD), P30 AG072931 (PI Henry Paulson, MD, PhD), P30 AG066546 (PI Sudha Seshadri, MD), P30 AG086401 (PI Erik Roberson, MD, PhD), P30 AG086404 (PI Gary Rosenberg, MD), P20 AG068082 (PI Angela Jefferson, PhD), P30 AG072958 (PI Heather Whitson, MD), P30 AG072959 (PI James Leverenz, MD).

For the purpose of open access, the author has applied a Creative Commons Attribution (CC BY) licence to any Author Accepted Manuscript version arising from this submission.

---

**Supplementary Table 1.** Mean cortical thickness and grey matter volume *Z*-score differences between no-NPS and MBI-apathy groups.

|  | No-NPS<br>(n=387) | MBI-apathy<br>(n=59) | *p*-value |
| --- | --- | --- | --- |
| **AD thickness meta-ROI** |  |  |  |
| Mean (SD) | -0.28 (0.92) | -0.90 (1.25) | **<0.001<sup>a</sup>** |
| Median [Min, Max] | -0.19 [-3.70, 1.19] | -0.63 [-3.70, 1.19] |  |
| **ACC thickness** |  |  |  |
| Mean (SD) | -0.11 (0.88) | -0.09 (1.17) | 0.856<sup>a</sup> |
| Median [Min, Max] | -0.08 [-2.39, 1.86] | 0.02 [-2.39, 1.86] |  |
| **OFC thickness** |  |  |  |
| Mean (SD) | -0.36 (0.96) | -0.67 (1.28) | **0.025<sup>a</sup>** |
| Median [Min, Max] | -0.38 [-3.77, 1.46] | -0.57 [-3.77, 1.46] |  |
| **vlPRF thickness** |  |  |  |
| Mean (SD) | -0.28 (0.85) | -0.49 (1.05) | 0.088<sup>a</sup> |
| Median [Min, Max] | -0.27 [-2.40, 1.41] | -0.45 [-2.40, 1.41] |  |
| **dlPFC thickness** |  |  |  |
| Mean (SD) | -0.26 (0.92) | -0.72 (1.19) | **0.001<sup>a</sup>** |
| Median [Min, Max] | -0.23 [-3.29, 1.33] | -0.67 [3.29, 1.33] |  |
| **AD volume meta-ROI** |  |  |  |
| Mean (SD) | -0.11 (0.62) | -0.79 (0.81) | **<0.001<sup>a</sup>** |
| Median [Min, Max] | -0.04 [-2.07, 1.01] | -0.83 [-2.07, 0.80] |  |
| **VS volume** |  |  |  |
| Mean (SD) | -0.03 (0.82) | -0.33 (0.97) | **0.012<sup>a</sup>** |
| Median [Min, Max] | -0.01 [-1.76, 1.67] | -0.25 [-1.76, 1.67] |  |
| **ACC volume** |  |  |  |
| Mean (SD) | 0.06 (0.65) | -0.06 (0.67) | 0.216<sup>a</sup> |
| Median [Min, Max] | 0.05 [-1.47, 1.35] | 0.03 [-1.47, 1.35] |  |
| **OFC volume** |  |  |  |
| Mean (SD) | 0.02 (0.80) | -0.47 (0.89) | **<0.001<sup>a</sup>** |
| Median [Min, Max] | 0.04 [-1.83, 1.55] | -0.57 [-1.83, 1.55] |  |
| **vlPRF volume** |  |  |  |
| Mean (SD) | -0.09 (0.66) | -0.21 (0.62) | 0.203<sup>a</sup> |
| Median [Min, Max] | -0.10 [-1.46, 1.32] | -0.20 [-1.46, 1.32] |  |
| **dlPFC volume** |  |  |  |
| Mean (SD) | -0.07 (0.69) | -0.45 (0.74) | **<0.001<sup>a</sup>** |
| Median [Min, Max] | -0.01 [-1.94, 1.17] | -0.48 [-1.94, 0.89] |  |

NPS = Neuropsychiatric Symptoms; MBI = Mild Behavioural Impairment; VS = Ventral striatum; ACC = Anterior cingulate cortex; OFC = Orbitofrontal cortex; vlPFC = Ventrolateral prefrontal cortex; dlPFC = Dorsolateral prefrontal cortex; ROI = Region of interest; SD = Standard deviation.
Significant *p*-values are shown in bold.
<sup>a</sup>Independent samples *t*-tests.

## References

1. Leung DKY, Chan WC, Spector A, Wong GHY. Prevalence of depression, anxiety, and apathy symptoms across dementia stages: A systematic review and meta-analysis. Int J Geriatr Psychiatry. Sep 2021;36(9):1330–1344. doi:10.1002/gps.5556

2. Pan Y, Shea YF, Ismail Z, et al. Prevalence of mild behavioural impairment domains: a meta-analysis. Psychogeriatrics. Jan 2022;22(1):84–98. doi:10.1111/psyg.12782

3. Marin RS. Differential diagnosis and classification of apathy. Am J Psychiatry. Jan 1990;147(1):22–30. doi:10.1176/ajp.147.1.22

4. Marin RS. Apathy: a neuropsychiatric syndrome. J Neuropsychiatry Clin Neurosci. Summer 1991;3(3):243–54. doi:10.1176/jnp.3.3.243

5. Miller DS, Robert P, Ereshefsky L, et al. Diagnostic criteria for apathy in neurocognitive disorders. Alzheimers Dement. Dec 2021;17(12):1892–1904. doi:10.1002/alz.12358

6. Chong TT. Definition: Apathy. Cortex. Jul 2020;128:326–327. doi:10.1016/j.cortex.2020.04.001

7. Ismail Z, Smith EE, Geda Y, et al. Neuropsychiatric symptoms as early manifestations of emergent dementia: Provisional diagnostic criteria for mild behavioral impairment. Alzheimers Dement. Feb 2016;12(2):195–202. doi:10.1016/j.jalz.2015.05.017

8. Ismail Z, Aguera-Ortiz L, Brodaty H, et al. The Mild Behavioral Impairment Checklist (MBI-C): A Rating Scale for Neuropsychiatric Symptoms in Pre-Dementia Populations. J Alzheimers Dis. 2017;56(3):929–938. doi:10.3233/JAD-160979

9. Hu S, Patten S, Charlton A, et al. Validating the Mild Behavioral Impairment Checklist in a Cognitive Clinic: Comparisons With the Neuropsychiatric Inventory Questionnaire. J Geriatr Psychiatry Neurol. Apr 17 2022:8919887221093353. doi:10.1177/08919887221093353

10. Creese B, Ismail Z. Mild behavioral impairment: measurement and clinical correlates of a novel marker of preclinical Alzheimer’s disease. Alzheimers Res Ther. Jan 5 2022;14(1):2. doi:10.1186/s13195-021-00949-7

11. Yoon EJ, Lee JY, Kwak S, Kim YK. Mild behavioral impairment linked to progression to Alzheimer’s disease and cortical thinning in amnestic mild cognitive impairment. Front Aging Neurosci. 2022;14:1051621. doi:10.3389/fnagi.2022.1051621

12. Guan DX, Rehman T, Nathan S, et al. Neuropsychiatric symptoms: Risk factor or disease marker? A study of structural imaging biomarkers of Alzheimer’s disease and incident cognitive decline. Hum Brain Mapp. Sep 2024;45(13):e70016. doi:10.1002/hbm.70016

13. Vellone D, Ghahremani M, Goodarzi Z, Forkert ND, Smith EE, Ismail Z. Apathy and APOE in mild behavioral impairment, and risk for incident dementia. Alzheimer’s Dement: Transl Res Clin Interv. 2022;8(1):1–12. doi:10.1002/trc2.12370

14. Vellone D, Leon R, Goodarzi Z, Forkert ND, Smith EE, Ismail Z. Mild behavioural impairment-apathy and core Alzheimer’s disease cerebrospinal fluid biomarkers. Brain. Jun 2 2025;doi:10.1093/brain/awaf194

15. Vellone D, Ismail Z. Associations Between Mild Behavioural Impairment-Apathy and Plasma p-Tau Levels. Can Geriatr J. 2025;28(3):292–293. doi:10.5770/cgj.28.884

16. Chan NK, Gerretsen P, Chakravarty MM, et al. Structural Brain Differences Between Cognitively Impaired Patients With and Without Apathy. Am J Geriatr Psychiatry. Apr 2021;29(4):319–332. doi:10.1016/j.jagp.2020.12.008

17. Yu SY, Zhu WL, Guo P, et al. Clinical features and brain structural changes in magnetic resonance imaging in Alzheimer’s disease patients with apathy. Aging (Albany NY). Oct 11 2020;12(19):19083–19094. doi:10.18632/aging.103705

18. Chen Y, Dang M, Zhang Z. Brain mechanisms underlying neuropsychiatric symptoms in Alzheimer’s disease: a systematic review of symptom-general and -specific lesion patterns. Mol Neurodegener. Jun 7 2021;16(1):38. doi:10.1186/s13024-021-00456-1

19. Stella F, Radanovic M, Aprahamian I, Canineu PR, de Andrade LP, Forlenza OV. Neurobiological correlates of apathy in Alzheimer’s disease and mild cognitive impairment: a critical review. J Alzheimers Dis. 2014;39(3):633–48. doi:10.3233/jad-131385

20. Mehak SF, Shivakumar AB, Saraf V, Johansson M, Gangadharan G. Apathy in Alzheimer’s disease: A neurocircuitry based perspective. Ageing Res Rev. Jun 2023;87:101891. doi:10.1016/j.arr.2023.101891

21. Theleritis C, Politis A, Siarkos K, Lyketsos CG. A review of neuroimaging findings of apathy in Alzheimer’s disease. Int Psychogeriatr. Feb 2014;26(2):195–207. doi:10.1017/S1041610213001725

22. Ghahremani M, Nathan S, Smith EE, McGirr A, Goodyear B, Ismail Z. Functional connectivity and mild behavioral impairment in dementia-free elderly. Alzheimers Dement (N Y). 2023;9(1):e12371. doi:10.1002/trc2.12371

23. Bock MA, Bahorik A, Brenowitz WD, Yaffe K. Apathy and risk of probable incident dementia among community-dwelling older adults. Neurology. Dec 15 2020;95(24):e3280–e3287. doi:10.1212/WNL.0000000000010951

24. Beekly DL, Ramos EM, van Belle G, et al. The National Alzheimer’s Coordinating Center (NACC) Database: an Alzheimer disease database. Alzheimer Dis Assoc Disord. Oct-Dec 2004;18(4):270–7.

25. Beekly DL, Ramos EM, Lee WW, et al. The National Alzheimer’s Coordinating Center (NACC) database: the Uniform Data Set. Alzheimer Dis Assoc Disord. Jul-Sep 2007;21(3):249–58. doi:10.1097/WAD.0b013e318142774e

26. Morris JC, Weintraub S, Chui HC, et al. The Uniform Data Set (UDS): clinical and cognitive variables and descriptive data from Alzheimer Disease Centers. Alzheimer Dis Assoc Disord. Oct-Dec 2006;20(4):210–6. doi:10.1097/01.wad.0000213865.09806.92

27. Besser L, Kukull W, Knopman DS, et al. Version 3 of the National Alzheimer’s Coordinating Center’s Uniform Data Set. Alzheimer Dis Assoc Disord. Oct-Dec 2018;32(4):351–358. doi:10.1097/wad.0000000000000279

28. Cummings J. The Neuropsychiatric Inventory: Development and Applications. J Geriatr Psychiatry Neurol. Mar 2020;33(2):73–84. doi:10.1177/0891988719882102

29. Guan DX, Smith EE, Pike BG, Ismail Z. Persistence of neuropsychiatric symptoms and dementia prognostication: A comparison of three operational case definitions of mild behavioral impairment. Alzheimers Dementia. 2023;15(4):e12483.

30. Cummings JL. The Neuropsychiatric Inventory–Questionnaire: Background and Administration. 1994:1–6.

31. Fischl B. FreeSurfer. Neuroimage. Aug 15 2012;62(2):774–81. doi:10.1016/j.neuroimage.2012.01.021

32. Fischl B, Salat DH, Busa E, et al. Whole brain segmentation: automated labeling of neuroanatomical structures in the human brain. Neuron. Jan 31 2002;33(3):341–55. doi:10.1016/s0896-6273(02)00569-x

33. Sherif T, Rioux P, Rousseau ME, et al. CBRAIN: a web-based, distributed computing platform for collaborative neuroimaging research. Front Neuroinform. 2014;8:54. doi:10.3389/fninf.2014.00054

34. Desikan RS, Ségonne F, Fischl B, et al. An automated labeling system for subdividing the human cerebral cortex on MRI scans into gyral based regions of interest. Neuroimage. Jul 1 2006;31(3):968–80. doi:10.1016/j.neuroimage.2006.01.021

35. Schwarz CG, Gunter JL, Wiste HJ, et al. A large-scale comparison of cortical thickness and volume methods for measuring Alzheimer’s disease severity. Neuroimage Clin. 2016;11:802–812. doi:10.1016/j.nicl.2016.05.017

36. Matuskova V, Ismail Z, Nikolai T, et al. Mild Behavioral Impairment Is Associated With Atrophy of Entorhinal Cortex and Hippocampus in a Memory Clinic Cohort. Front Aging Neurosci. 2021;13:643271. doi:10.3389/fnagi.2021.643271

37. Donovan NJ, Wadsworth LP, Lorius N, et al. Regional cortical thinning predicts worsening apathy and hallucinations across the Alzheimer disease spectrum. Am J Geriatr Psychiatry. Nov 2014;22(11):1168–79. doi:10.1016/j.jagp.2013.03.006

38. Guercio BJ, Donovan NJ, Ward A, et al. Apathy is associated with lower inferior temporal cortical thickness in mild cognitive impairment and normal elderly individuals. J Neuropsychiatry Clin Neurosci. Winter 2015;27(1):e22–7. doi:10.1176/appi.neuropsych.13060141

39. Zhao H, Tang W, Xu X, Zhao Z, Huang L. Functional Magnetic Resonance Imaging Study of Apathy in Alzheimer’s Disease. J Neuropsychiatry Clin Neurosci. Spring 2014;26(4):134–141. doi:10.1176/appi.neuropsych.12110261

40. Kang JY, Lee JS, Kang H, et al. Regional cerebral blood flow abnormalities associated with apathy and depression in Alzheimer disease. Alzheimer Dis Assoc Disord. Jul-Sep 2012;26(3):217–24. doi:10.1097/WAD.0b013e318231e5fc

41. Robert PH, Darcourt G, Koulibaly MP, et al. Lack of initiative and interest in Alzheimer’s disease: a single photon emission computed tomography study. Eur J Neurol. Jul 2006;13(7):729–35. doi:10.1111/j.1468-1331.2006.01088.x

42. Premnath PY, Locascio JJ, Mimmack KJ, et al. Longitudinal associations of apathy and regional tau in mild cognitive impairment and dementia: Findings from the Alzheimer’s Disease Neuroimaging Initiative. Alzheimers Dement (N Y). Jan-Mar 2024;10(1):e12442. doi:10.1002/trc2.12442

43. Joo SH, Lee CU, Lim HK. Apathy and intrinsic functional connectivity networks in amnestic mild cognitive impairment. Neuropsychiatr Dis Treat. 2017;13:61–67. doi:10.2147/ndt.S123338

44. Katz ZS, Mimmack KJ, Marshall GA, et al. Association of Apathy with Amyloid-β and Tau in Cognitively Unimpaired Older Adults: Findings from the Harvard Aging Brain Study. Alzheimer’s & Dementia. 2022;18(S7):e067122. 10.1002/alz.067122

45. Johansson M, Stomrud E, Insel PS, et al. Mild behavioral impairment and its relation to tau pathology in preclinical Alzheimer’s disease. Transl Psychiatry. Jan 26 2021;11(1):76. doi:10.1038/s41398-021-01206-z

46. Taragano FE, Allegri RF, Krupitzki H, et al. Mild behavioral impairment and risk of dementia: a prospective cohort study of 358 patients. J Clin Psychiatry. Apr 2009;70(4):584–92. doi:10.4088/jcp.08m04181

47. Lussier F. Alzheimer’s disease pathophysiological correlates of the syndrome of mild behavioral impairment across the spectrum of the disease. Thesis. McGill University; 2022. https://escholarship.mcgill.ca/concern/theses/r494vq95q

48. Braak H, Braak E. Staging of Alzheimer’s disease-related neurofibrillary changes. Neurobiol Aging. May-Jun 1995;16(3):271–8; discussion 278-84. doi:10.1016/0197-4580(95)00021-6

49. Braak H, Alafuzoff I, Arzberger T, Kretzschmar H, Del Tredici K. Staging of Alzheimer disease-associated neurofibrillary pathology using paraffin sections and immunocytochemistry. Acta Neuropathol. Oct 2006;112(4):389–404. doi:10.1007/s00401-006-0127-z

50. Apostolova LG, Akopyan GG, Partiali N, et al. Structural correlates of apathy in Alzheimer’s disease. Dement Geriatr Cogn Disord. 2007;24(2):91–7. doi:10.1159/000103914

51. Bruen PD, McGeown WJ, Shanks MF, Venneri A. Neuroanatomical correlates of neuropsychiatric symptoms in Alzheimer’s disease. Brain. Sep 2008;131(Pt 9):2455–63. doi:10.1093/brain/awn151

52. Tunnard C, Whitehead D, Hurt C, et al. Apathy and cortical atrophy in Alzheimer’s disease. Int J Geriatr Psychiatry. Jul 2011;26(7):741–8. doi:10.1002/gps.2603

53. Huey ED, Lee S, Cheran G, Grafman J, Devanand DP, Alzheimer’s Disease Neuroimaging I. Brain Regions Involved in Arousal and Reward Processing are Associated with Apathy in Alzheimer’s Disease and Frontotemporal Dementia. J Alzheimers Dis. 2017;55(2):551–558. doi:10.3233/JAD-160107

54. de Jong LW, Wang Y, White LR, Yu B, van Buchem MA, Launer LJ. Ventral striatal volume is associated with cognitive decline in older people: a population based MR-study. Neurobiol Aging. Feb 2012;33(2):424.e1-10. doi:10.1016/j.neurobiolaging.2010.09.027

55. Stanton BR, Leigh PN, Howard RJ, Barker GJ, Brown RG. Behavioural and emotional symptoms of apathy are associated with distinct patterns of brain atrophy in neurodegenerative disorders. J Neurol. Oct 2013;260(10):2481–90. doi:10.1007/s00415-013-6989-9

56. Mohamed Nour AEA, Jiao Y, Teng GJ, Alzheimer’s Disease Neuroimaging I. Neuroanatomical associations of depression, anxiety and apathy neuropsychiatric symptoms in patients with Alzheimer’s disease. Acta Neurol Belg. Dec 2021;121(6):1469–1480. doi:10.1007/s13760-020-01349-8

57. Moon Y, Moon WJ, Kim H, Han SH. Regional atrophy of the insular cortex is associated with neuropsychiatric symptoms in Alzheimer’s disease patients. Eur Neurol. 2014;71(5-6):223–9. doi:10.1159/000356343

58. Le Heron C, Holroyd CB, Salamone J, Husain M. Brain mechanisms underlying apathy. J Neurol Neurosurg Psychiatry. Mar 2019;90(3):302–312. doi:10.1136/jnnp-2018-318265

59. Kim JW, Lee DY, Choo IH, et al. Microstructural alteration of the anterior cingulum is associated with apathy in Alzheimer disease. Am J Geriatr Psychiatry. Jul 2011;19(7):644–53. doi:10.1097/JGP.0b013e31820dcc73

60. Tighe SK, Oishi K, Mori S, et al. Diffusion tensor imaging of neuropsychiatric symptoms in mild cognitive impairment and Alzheimer’s dementia. J Neuropsychiatry Clin Neurosci. Fall 2012;24(4):484–8. doi:10.1176/appi.neuropsych.11120375

61. Ota M, Sato N, Nakata Y, Arima K, Uno M. Relationship between apathy and diffusion tensor imaging metrics of the brain in Alzheimer’s disease. Int J Geriatr Psychiatry. Jul 2012;27(7):722–6. doi:10.1002/gps.2779

62. Totuk O, Sahin S. Apathy in Dementia: A Pilot Study Providing Insights from Neuropsychiatric and Radiological Perspectives. J Clin Med. Mar 8 2025;14(6)doi:10.3390/jcm14061822

63. Starkstein SE, Mizrahi R, Capizzano AA, Acion L, Brockman S, Power BD. Neuroimaging correlates of apathy and depression in Alzheimer’s disease. J Neuropsychiatry Clin Neurosci. Summer 2009;21(3):259–65. doi:10.1176/jnp.2009.21.3.259

64. Büyükgök D, Bayraktaroğlu Z, Buker HS, Kulaksızoğlu MIB, Gurvit İ H. Resting-state fMRI analysis in apathetic Alzheimer’s disease. Diagn Interv Radiol. Jul 2020;26(4):363–369. doi:10.5152/dir.2019.19445

65. Huang SM, Hsu YH, Yang JJ, Lin CY, Tu MC, Kuo LW. Functional and microstructural neurosubstrates between apathy and depressive symptoms in dementia. Neuroimage Clin. 2025;46:103781. doi:10.1016/j.nicl.2025.103781

66. Jones SA, De Marco M, Manca R, et al. Altered frontal and insular functional connectivity as pivotal mechanisms for apathy in Alzheimer’s disease. Cortex. Oct 2019;119:100–110. doi:10.1016/j.cortex.2019.04.008

67. Migneco O, Benoit M, Koulibaly PM, et al. Perfusion brain SPECT and statistical parametric mapping analysis indicate that apathy is a cingulate syndrome: a study in Alzheimer’s disease and nondemented patients. Neuroimage. May 2001;13(5):896–902. doi:10.1006/nimg.2000.0741

68. Benoit M, Koulibaly PM, Migneco O, Darcourt J, Pringuey DJ, Robert PH. Brain perfusion in Alzheimer’s disease with and without apathy: a SPECT study with statistical parametric mapping analysis. Psychiatry Res. Jun 15 2002;114(2):103–11. doi:10.1016/s0925-4927(02)00003-3

69. Lanctôt KL, Moosa S, Herrmann N, et al. A SPECT study of apathy in Alzheimer’s disease. Dement Geriatr Cogn Disord. 2007;24(1):65–72. doi:10.1159/000103633

70. Valotassiou V, Sifakis N, Tzavara C, et al. Differences of apathy perfusion correlates between Alzheimer’s disease and frontotemporal dementia. A 99mTc-HMPAO SPECT study with automated Brodmann areas analysis. Int J Psychiatry Clin Pract. Mar 2022;26(1):14–22. doi:10.1080/13651501.2020.1846752

71. Craig AH, Cummings JL, Fairbanks L, et al. Cerebral blood flow correlates of apathy in Alzheimer disease. Arch Neurol. Nov 1996;53(11):1116–20. doi:10.1001/archneur.1996.00550110056012

72. Starkstein SE, Sabe L, Vázquez S, et al. Neuropsychological, psychiatric, and cerebral perfusion correlates of leukoaraiosis in Alzheimer’s disease. J Neurol Neurosurg Psychiatry. Jul 1997;63(1):66–73. doi:10.1136/jnnp.63.1.66

73. Benoit M, Dygai I, Migneco O, et al. Behavioral and psychological symptoms in Alzheimer’s disease. Relation between apathy and regional cerebral perfusion. Dement Geriatr Cogn Disord. Nov-Dec 1999;10(6):511–7. doi:10.1159/000017198

74. Marshall GA, Fairbanks LA, Tekin S, Vinters HV, Cummings JL. Neuropathologic correlates of apathy in Alzheimer’s disease. Dement Geriatr Cogn Disord. 2006;21(3):144–7. doi:10.1159/000090674

75. Mori T, Shimada H, Shinotoh H, et al. Apathy correlates with prefrontal amyloid β deposition in Alzheimer’s disease. J Neurol Neurosurg Psychiatry. Apr 2014;85(4):449–55. doi:10.1136/jnnp-2013-306110

76. Benoit M, Clairet S, Koulibaly PM, Darcourt J, Robert PH. Brain perfusion correlates of the apathy inventory dimensions of Alzheimer’s disease. Int J Geriatr Psychiatry. Sep 2004;19(9):864–9. doi:10.1002/gps.1163

77. Marshall GA, Monserratt L, Harwood D, Mandelkern M, Cummings JL, Sultzer DL. Positron emission tomography metabolic correlates of apathy in Alzheimer disease. Arch Neurol. Jul 2007;64(7):1015–20. doi:10.1001/archneur.64.7.1015

78. Kitamura S, Shimada H, Niwa F, et al. Tau-induced focal neurotoxicity and network disruption related to apathy in Alzheimer’s disease. J Neurol Neurosurg Psychiatry. Nov 2018;89(11):1208–1214. doi:10.1136/jnnp-2018-317970

79. Holthoff VA, Beuthien-Baumann B, Kalbe E, et al. Regional cerebral metabolism in early Alzheimer’s disease with clinically significant apathy or depression. Biol Psychiatry. Feb 15 2005;57(4):412–21. doi:10.1016/j.biopsych.2004.11.035

80. Mega MS, Dinov ID, Porter V, et al. Metabolic patterns associated with the clinical response to galantamine therapy: a fludeoxyglucose f 18 positron emission tomographic study. Arch Neurol. May 2005;62(5):721–8. doi:10.1001/archneur.62.5.721

81. Schroeter ML, Vogt B, Frisch S, et al. Dissociating behavioral disorders in early dementia-An FDG-PET study. Psychiatry Res. Dec 30 2011;194(3):235–244. doi:10.1016/j.pscychresns.2011.06.009

82. Grool AM, Geerlings MI, Sigurdsson S, et al. Structural MRI correlates of apathy symptoms in older persons without dementia: AGES-Reykjavik Study. Neurology. May 6 2014;82(18):1628–35. doi:10.1212/wnl.0000000000000378

83. Johansson M, Stomrud E, Lindberg O, et al. Apathy and anxiety are early markers of Alzheimer’s disease. Neurobiol Aging. Jan 2020;85:74–82. doi:10.1016/j.neurobiolaging.2019.10.008

84. Yan H, Onoda K, Yamaguchi S. Gray Matter Volume Changes in the Apathetic Elderly. Front Hum Neurosci. 2015;9:318. doi:10.3389/fnhum.2015.00318

85. Torso M, Serra L, Giulietti G, et al. Strategic lesions in the anterior thalamic radiation and apathy in early Alzheimer’s disease. PLoS One. 2015;10(5):e0124998. doi:10.1371/journal.pone.0124998

86. Marshall GA, Donovan NJ, Lorius N, et al. Apathy is associated with increased amyloid burden in mild cognitive impairment. J Neuropsychiatry Clin Neurosci. Fall 2013;25(4):302–7. doi:10.1176/appi.neuropsych.12060156

87. Krell-Roesch J, Ruider H, Lowe VJ, et al. FDG-PET and Neuropsychiatric Symptoms among Cognitively Normal Elderly Persons: The Mayo Clinic Study of Aging. J Alzheimers Dis. Jul 14 2016;53(4):1609–16. doi:10.3233/jad-160326

88. Zahodne LB, Gongvatana A, Cohen RA, Ott BR, Tremont G. Are apathy and depression independently associated with longitudinal trajectories of cortical atrophy in mild cognitive impairment? Am J Geriatr Psychiatry. Nov 2013;21(11):1098–106. doi:10.1016/j.jagp.2013.01.043

89. Orso B, Mattei C, Arnaldi D, et al. Clinical and MRI Predictors of Conversion From Mild Behavioural Impairment to Dementia. Am J Geriatr Psychiatry. Jul 2020;28(7):755–763. doi:10.1016/j.jagp.2019.12.007

90. Shu J, Qiang Q, Yan Y, et al. Distinct Patterns of Brain Atrophy associated with Mild Behavioral Impairment in Cognitively Normal Elderly Adults. Int J Med Sci. 2021;18(13):2950–2956. doi:10.7150/ijms.60810

91. Shu J, Qiang Q, Yan Y, Ren Y, Wei W, Zhang L. Aberrant Topological Patterns of Structural Covariance Networks in Cognitively Normal Elderly Adults With Mild Behavioral Impairment. Front Neuroanat. 2021;15:738100. doi:10.3389/fnana.2021.738100

92. Yang L, Shu J, Yan A, Yang F, Xu Z, Wei W. White matter hyperintensities-related cortical changes and correlation with mild behavioral impairment. Adv Med Sci. Sep 2022;67(2):241–249. doi:10.1016/j.advms.2022.06.002

93. Matsuoka T, Ueno D, Ismail Z, et al. Neural Correlates of Mild Behavioral Impairment: A Functional Brain Connectivity Study Using Resting-State Functional Magnetic Resonance Imaging. J Alzheimers Dis. 2021;83(3):1221–1231. doi:10.3233/jad-210628

94. Levy R, Dubois B. Apathy and the functional anatomy of the prefrontal cortex-basal ganglia circuits. Cereb Cortex. Jul 2006;16(7):916–28. doi:10.1093/cercor/bhj043

95. Gatchel JR, Donovan NJ, Locascio JJ, et al. Regional 18F-Fluorodeoxyglucose Hypometabolism is Associated with Higher Apathy Scores Over Time in Early Alzheimer Disease. Am J Geriatr Psychiatry. Jul 2017;25(7):683–693. doi:10.1016/j.jagp.2016.12.017

96. Chow TW, Binns MA, Cummings JL, et al. Apathy symptom profile and behavioral associations in frontotemporal dementia vs dementia of Alzheimer type. Arch Neurol. Jul 2009;66(7):888–93. doi:10.1001/archneurol.2009.92

97. Vasconcelos Da Silva M, Melendez-Torres GJ, Ismail Z, Testad I, Ballard C, Creese B. A data-driven examination of apathy and depressive symptoms in dementia with independent replication. Alzheimers Dement (Amst). Jan-Mar 2023;15(1):e12398. doi:10.1002/dad2.12398

98. Valles KJ, Ayers E, Verghese J, Ceïde ME. Diminished Social and Leisure Engagement in Community Dwelling-Older Adults with Apathy. Int J Environ Res Public Health. Jul 18 2025;22(7)doi:10.3390/ijerph22071138

99. Harrison F, Mortby ME, Mather KA, Sachdev PS, Brodaty H. Apathy as a determinant of health behaviors in older adults: Implications for dementia risk reduction. Alzheimers Dement (Amst). Oct-Dec 2023;15(4):e12505. doi:10.1002/dad2.12505

100. Guan DX, Mortby ME, Pike GB, et al. Linking cognitive and behavioral reserve: Evidence from the CAN-PROTECT study. Alzheimers Dement (N Y). Oct 2024;10(4):e12497. doi:10.1002/trc2.12497

101. Mudalige D, Guan DX, Ballard C, et al. The mind and motion: exploring the interplay between physical activity and Mild Behavioral Impairment in dementia-free older adults. Int Rev Psychiatry. May 2024;36(3):196–207. doi:10.1080/09540261.2024.2360561

